# Early Estimation Of Reproduction Number of Covid-19 in Vietnam

**DOI:** 10.1101/2020.03.28.20046136

**Authors:** Long Bui, Truong Nguyen Thanh, Ha Nguyen Ngoc

## Abstract

Reproduction number is an epidemiologic indicator that reflects the contagiousness and transmissibility of infectious agents. This paper aims to estimate the reproduction number of in the early phase of COVID-19 outbreak in Vietnam.

## Introduction

Reproduction number is an epidemiologic indicator that reflects the contagiousness and transmissibility of infectious agents. Understanding the reproduction number is crucial for predicting the transmission of an infectious diseases and evaluating the effectiveness of control measures.

There are several ways to estimate the reproduction number, including mathematical modeling of compartment models or through the serial interval of the infection chain [1,2]. The estimation of reproduction number by compartment models is heavily based on several assumptions, such as a homogeneous mixing of the fixed population [3]. The parameters, including incubation period, and infectious period are often not available with novel infectious disease line COVID-19. Alternatively, estimation of reproduction number by the mean and standard deviation of serial interval, which is the range between primary and secondary symptom onset dates is an uncomplicated approach.

## Methods

We collected data on confirmed Covid-19 cases in Vietnam from multiple sources. The master list of Covid-19 cases was compiled from the official new releases of Ministry of Health (available on the website http://ncov.moh.ncov.vn). The first two Covid-19 cases were described by Lan et al. [4]. Latterly, 11 confirmed cases from one northern Vietnam province were published [5]. Epidemiologic history from data was combined with the master list.

We construct a network and make plausible cases of infectors and infectees, and there by estimate the discrete distribution of serial interval from the chain of infections. We assumed that the serial interval follows the discrete gamma distribution. The reproduction number was estimate by the growth rate of the epidemiological curve and the mean and standard deviation (SD) of the gamma distribution. We used R 3.6.4 software [5] for data analysis and modeling. Package *incidence* [6] was used to create incidence object. The serial interval distribution and early reproduction number was estimated by package *earlyR* [7].

## Results

From 20 Jan 2020 to 24 March 2020, there was 123 cases confirmed with positive SARS-CoV-2 was reported in Vietnam. The epidemiologic trend of Covid-19 relatively divided into two periods Figure 1. The first period started from 17 Jan 20 to 11 Feb 2020. In this period, 9 of 16 confirmed cases were imported. The network of these 16 cases and chain of infections was constructed (Table 1). We estimated the mean of discrete gamma distribution of serial interval was 5.83 and SD was 3.58.

**Table 1.** Estimated serial interval of COVID-19 from chain of infection.

| From | To | Onset From | Onset To | Serial Interval |
| --- | --- | --- | --- | --- |
| Patient 1 | Patient 2 | 17/01/2020 | 20/01/2020 | 4 |
| Patient 1 | Patient 6 | 17/01/2020 | 18/01/2020 | 2 |
| Patient 5 | Patient 11 | 26/01/2020 | 03/02/2020 | 8 |
| Patient 5 | Patient 12 | 26/01/2020 | 06/02/2020 | 11 |
| Patient 5 | Patient 10 | 26/01/2020 | 01/02/2020 | 7 |
| Patient 10 | Patient 15 | 03/02/2020 | 11/02/2020 | 8 |
| Patient 24 | Patient 38 | 07/02/2020 | 08/03/2020 | 1 |
| Patient 86 | Patient 87 | 11/03/2020 | 18/03/2020 | 7 |
| Patient 91 | Patient 120 | 16/03/2020 | 21/03/2020 | 4 |

**Figure 1.**
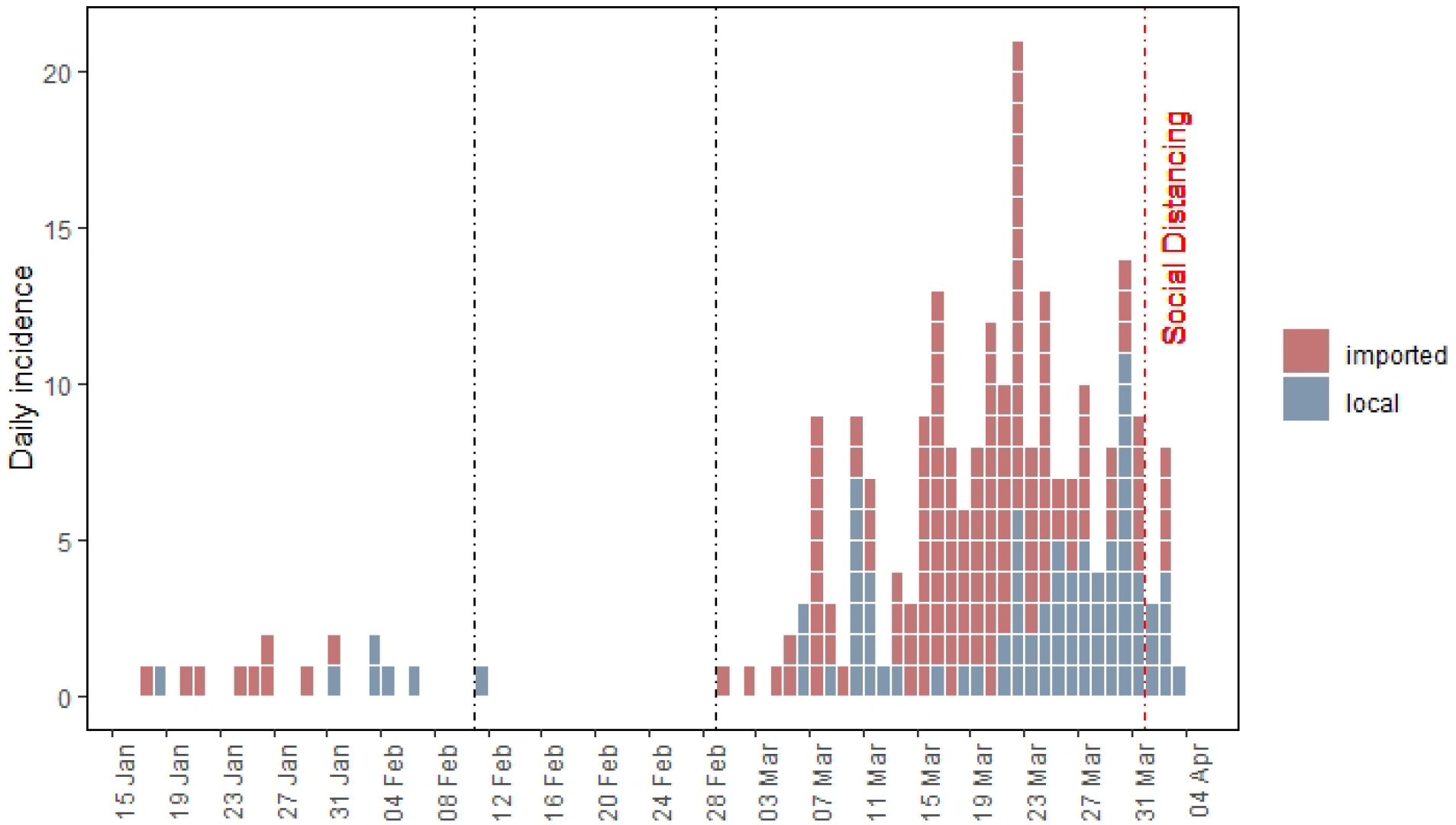
Epidemic curve of COVID-19 in Vietnam (From 17 Jan 2020 to 23 Mar 2020)

**Figure 2.**
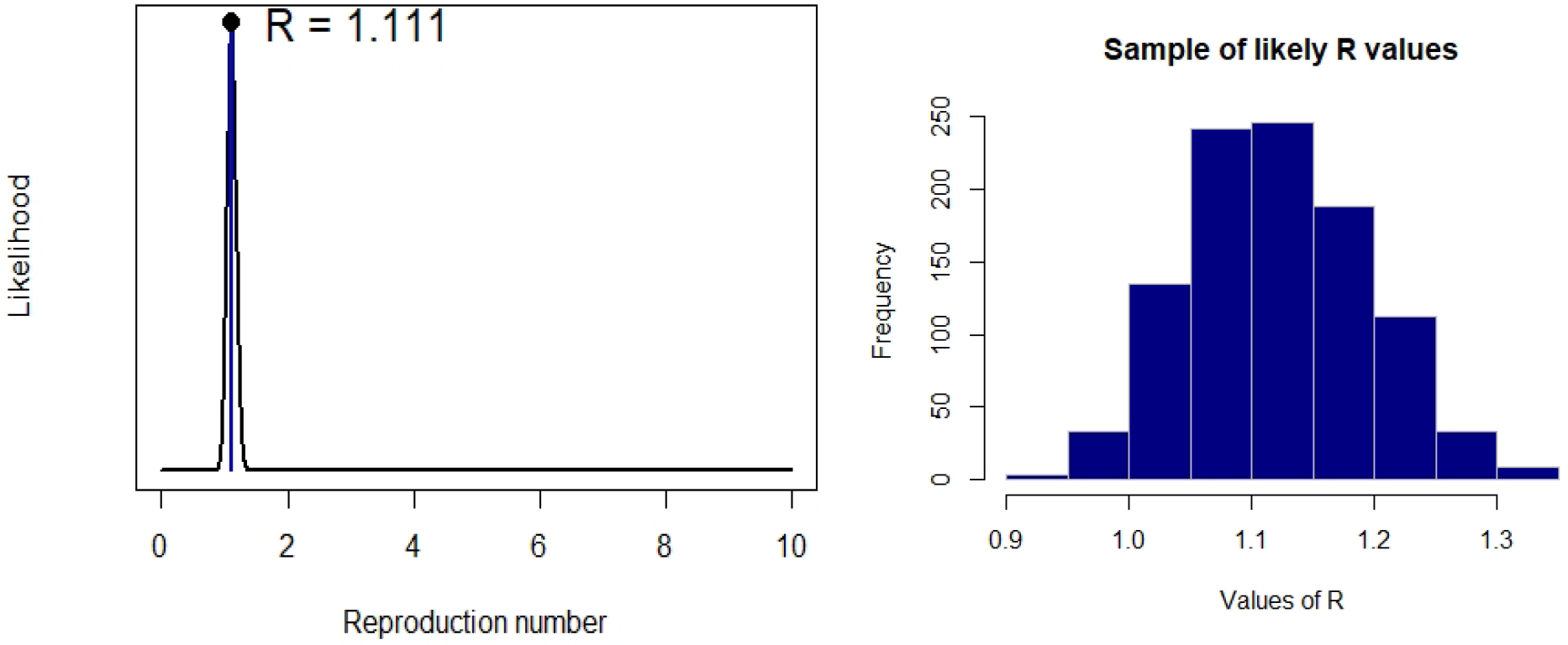
Estimated reproduction number of COVID-19 in Vietnam (From 17 Jan 2020 to 05 Apr 2020)

The mean estimated reproduction number of Covid-19 in Vietnam until 05 Apr Mar 2020 was 1.1 (median 1.11, 95%CI 0.98 – 1.26, 1000 bootstraps) (Figure 2)

## Discussion

Using publicly reported data, we applied a simple model to estimate the reproduction number of early stage of COVID-19 outbreak in Vietnam. The mean estimated reproduction number of Covid-19 in Vietnam until 05 Apr 2020 was 1.11. With the early response and active control measures of Vietnamese government, including mandatory isolating patients and suspected cases, the reproduction number of COVID-19 in Vietnam is considerable lower than reported in other countries [2].

It is worth noting that the epidemiologic data, including date of onset, date of positive-confirmed and contact tracing of first 16 COVID-19 cases in Vietnam were more detailed than latter cases. The fitted distribution of serial interval of this study is lower than described by Li el al. [9], which was also used by Zhang et al. to estimate the reproduction number on the Diamond Princess cruise ship [10].

Nevertheless, the analysis may shed lights on the reproduction number of COVID-19 outbreaks in Vietnam. The same methods can be applied with more detailed data on the chains of infection, and get better results and help policy makers monitor and evaluate the effectiveness of control measures.

## Data Availability

The data for analysis is available.

https://github.com/longbui/VN-COVID19

## Disclaimer

The views and opinions expressed in this article are those of the authors and do not necessarily reflect the official policy or position of any agency of the government.

## IRB and/or ethics committee approvals

Not Applicable

## References

1. Zhao S, Lin Q, Ran J, Musa SS, Yang G, Wang W, et al. Preliminary estimation of the basic reproduction number of novel coronavirus (2019-nCoV) in China, from 2019 to 2020: A data-driven analysis in the early phase of the outbreak. International Journal of Infectious Diseases. 2020 Mar 1;92:214–7.

2. Liu Y, Gayle AA, Wilder-Smith A, Rocklöv J. The reproductive number of COVID-19 is higher compared to SARS coronavirus. J Travel Med [Internet]. [cited 2020 Feb 16]; Available from: https://academic.oup.com/jtm/advance-article/doi/10.1093/jtm/taaa021/5735319

3. Sun C, Hsieh Y-H. Global analysis of an SEIR model with varying population size and vaccination. Applied Mathematical Modelling. 2010 Oct 1;34(10):2685–97.

4. Phan LT, Nguyen TV, Luong QC, Nguyen TV, Nguyen HT, L. Hq, et al. Importation and Human-to-Human Transmission of a Novel Coronavirus in Vietnam. New England Journal of Medicine [Internet]. 2020 Jan 28 [cited 2020 Feb 16]; Available from: https://doi.org/10.1056/NEJMc2001272

5. Outbreak investigation for COVID-19 in northern Vietnam - The Lancet Infectious Diseases [Internet]. [cited 2020 Mar 26]. Available from: https://www.thelancet.com/journals/laninf/article/PIIS1473-3099(20)30159-6/fulltext

6. Kamvar ZN, Cai J, Pulliam JRC, Schumacher J, Jombart T. Epidemic curves made easy using the R package incidence. F1000Res [Internet]. 2019 Jan 31 [cited 2020 Mar 26];8. Available from: https://www.ncbi.nlm.nih.gov/pmc/articles/PMC6509961/

7. Cori A, Ferguson NM, Fraser C, Cauchemez S. A New Framework and Software to Estimate Time-Varying Reproduction Numbers During Epidemics. Am J Epidemiol. 2013 Nov 1;178(9):1505–12.

8. Jombart T, Nouvellet P. projections: Project Future Case Incidence [Internet]. 2018. Available from: https://CRAN.R-project.org/package=projections

9. Li Q, Guan X, Wu P, Wang X, Zhou L, Tong Y, et al. Early Transmission Dynamics in Wuhan, China, of Novel Coronavirus–Infected Pneumonia. New England Journal of Medicine. 2020 Mar 26;382(13):1199–207.

10. Zhang S, Diao M, Yu W, Pei L, Lin Z, Chen D. Estimation of the reproductive number of novel coronavirus (COVID-19) and the probable outbreak size on the Diamond Princess cruise ship: A data-driven analysis. International Journal of Infectious Diseases. 2020 Apr 1;93:201–4.

